# Insights into Predicting Tooth Extraction from Panoramic Dental Images: Artificial Intelligence vs. Dentists

**DOI:** 10.1101/2024.04.22.24306189

**Authors:** Ila Motmaen, Kunpeng Xie, Leon Schönbrunn, Jeff Berens, Kim Grunert, Anna Maria Plum, Johannes Raufeisen, André Ferreira, Alexander Hermans, Jan Egger, Frank Hölzle, Daniel Truhn, Behrus Puladi

## Abstract

**Objectives:** Tooth extraction is one of the most frequently performed medical procedures. The indication is based on the combination of clinical and radiological examination and individual patient parameters and should be made with great care. However, determining whether a tooth should be extracted is not always a straightforward decision. Moreover, visual and cognitive pitfalls in the analysis of radiographs may lead to incorrect decisions. Artificial intelligence (AI) could be used as a decision support tool to provide a score of tooth extractability.

**Material and Methods:** Using 26,956 single teeth images from 1,184 panoramic radiographs (PANs), we trained a ResNet50 network to classify teeth as either extraction-worthy or preservable. For this purpose, teeth were cropped with different margins from PANs and annotated. The usefulness of the AI-based classification as well that of dentists was evaluated on a test dataset. In addition, the explainability of the best AI model was visualized via a class activation mapping using CAMERAS.

**Results:** The ROC-AUC for the best AI model to discriminate teeth worthy of preservation was 0.901 with 2% margin on dental images. In contrast, the average ROC-AUC for dentists was only 0.797. With a 19.1% tooth extractions prevalence, the AI model’s PR-AUC was 0.749, while the human evaluation only reached 0.589.

**Conclusion:** AI models outperform dentists/specialists in predicting tooth extraction based solely on X-ray images, while the AI performance improves with increasing contextual information.

**Clinical Relevance:** AI could help monitor at-risk teeth and reduce errors in indications for extractions.

## Introduction

Tooth extraction is one of the most commonly performed medical measures in the field of general dentistry/ oral and maxillofacial surgery. The decision is based on the patient’s records, which include medical history, clinical evaluation, and radiographs. Given its irreversible impact on the quality of life, the decision of extraction should be made with great care [1–3]. Certain X-ray signs are pivotal in determining the necessity for tooth extraction. These signs include the compromised structural integrity of the tooth, significant alveolar bone loss, or evident root fractures. In addition, massive periapical radiolucency may also suggest the extraction. Advanced internal or external resorption cases can also be identified on these radiographs, providing a clear indication for removal of the affected teeth [4].

Although indications are made clear in the extraction guidelines [5, 6], the decision-making process is not always easy for the practitioner in clinical practice [2, 4]. This decision may be confounded by many factors, such as the dentist’s/specialist’s own experience, the reliability of the clinical evidence, or even pressure from patients [5]. The interplay of these different potentially disruptive factors regarding diagnostic decision-making can lead to misdiagnosis and problematic therapy situations, especially in borderline cases. For example, incorrect tooth extraction is the third most common cause of tooth loss in periodontally damaged teeth [7].

However, leaving teeth that are not worthy of preservation is not an option, as they can cause massive pain [1] and can even be the starting point for life-threatening lodge abscesses in the head and neck region or cause fatal endocarditis, which ultimately affects the entire organism [8, 9]. At the same time, every tooth extraction has its risk of serious complications like persisting root fractures, dry sockets or damage to neighboring teeth. The indication is, therefore, also always a balancing of different requirements. In general, tooth extraction serves as a last resort when every other treatment option failed or is not indicated anymore [4].

Panoramic radiographs (PANs), commonly used due to easy access and low dosage, are crucial in evaluating a patient’s dental condition, providing insights into the whole dentition and relating structures [10]. However, accurate and comprehensive interpretation of PANs requires extensive training and considerable clinical experience. This expertise may not be fully developed in young practitioners, potentially leading to variability in diagnostic decisions [11]. Furthermore, seasoned practitioners may also be susceptible to cognitive and visual pitfalls when dealing with challenging cases [12].

Deep learning (DL), a subfield of artificial intelligence (AI), has revolutionized the field of medical imaging by extending the capabilities of human practitioners. These models are trained on vast datasets, allowing them to recognize patterns and anomalies with superhuman precision [13]. In the context of PANs, the DL models enable the detection and segmentation of anatomical structures in seconds, with performance improvements being noted on an ongoing basis [14–18]. Moreover, DL models can identify subtle or complex pathologies that may be overlooked by the human eye, such as caries, cysts, periodontitis, and periapical lesions. These can be automatically annotated with high accuracy [19–23]. Such advancements demonstrate the potential of DL to serve as a powerful tool that enhances diagnostic accuracy and efficiency.

Despite these advancements, most research has focused on lesion diagnosis [24–28], with limited exploration into subsequent clinical decisions like tooth extraction. Furthermore, the model’s predictions are often given with blunt probabilities without any explanation or reasoning process, which is crucial for clinical acceptance and understanding. Applying explainable DL has the potential to accelerate the decision-making process, resulting in timely and more effective interventions, ultimately leading to improved patient outcomes [29].

The study’s main objective is to develop and internally validate a model that can predict the need for tooth extraction from PANs and compare its performance to dentists/specialists. Furthermore, the effect of contextual knowledge of teeth on the model’s performance and its possible explainability will be visualized.

## Material and Methods

### Study Design and Patients

The study used retrospective PANs from 2011 to 2021 from patients who underwent tooth extraction at the Department of Oral and Maxillofacial Surgery of the University Hospital RWTH Aachen. Patients with edentulous conditions, or without available panoramic radiographs taken within six months post-treatment were excluded. Additionally, patients with significant artifacts in their preoperative panoramic radiographs that affected the teeth were also removed from the study cohort.

The study was approved by the Ethics Committee of the University Hospital RWTH Aachen (approval number EK 068/21, chairs: Prof. Dr. G. Schmalzing and PD Dr. R. Hausmann, approval date 25.02.2021) and followed the MI-CLAIM reporting guideline for the development of AI models [30].

### Dataset Preparation

For the study, all PANs were exported in DICOM format from the hospital’s picture archiving and communication system. If a patient had received more than one PAN within six months post-treatment, the last PAN would be taken as the postoperative image. After the cohort’s statistical summary, all PANs were stratified by patients and converted to PNG format for anonymization purposes.

Annotations and labeling of teeth in the preoperative PANs were performed by four investigators (I.M., J.B., K.G. and B.P.) using LabelMe [31]. For this purpose, all teeth were marked with a bounding box on the preoperative image and divided into a preserved and extracted class according to their presence in the postoperative image (Figure 1). Implants or residual roots were marked in the same way as teeth. For quality control, the annotated images and labels were then reviewed by two investigators (I.M. and B.P.) for a second round.

**Figure 1.**
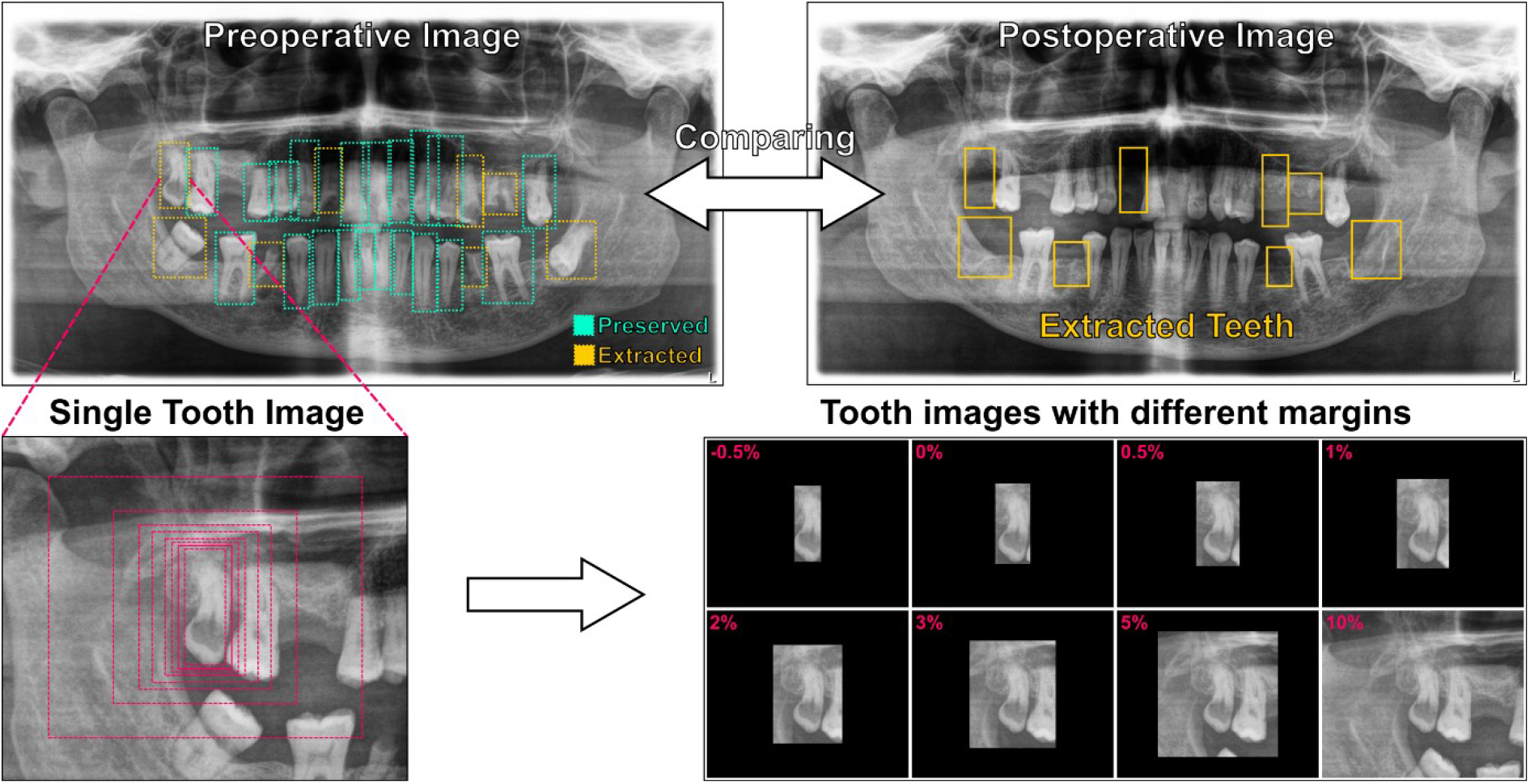
Pipeline to prepare the dataset. Panoramic radiographs from the same patient were compared and annotations of teeth were made on the preoperative image with bounding boxes and labeled as preserved (green) or extracted (yellow). Different margin factors were used to resize the bounding boxes (red) in width and height. Teeth images were then cropped from the original image with margins (-0.5% to 10%).

The bounding boxes were then used to export single tooth images with different margins, as well as their class (preserved or extracted tooth). Since the distances (in mm) in PANs are not uniform and the teeth themselves have different sizes, we defined the margins in % of the PAN image height and width. Images were then exported with margins ranging from -0.5% to 10%, with 0% being the bounding box itself, resulting in 8 datasets. Figure 1 describes the pipeline of the dataset preparation.

### Model Development and Validation

The dataset was stratified by patient and randomly divided into a training set (17,874), validation set (4,784), and test set (4,298) in a 4:1:1 ratio. During training, we apply a random crop to the image, then resize it to 224x224 pixels and perform horizontal flip augmentations to enhance model generalization. Validation and test sets images are resized to 256x256 pixels and the 224x224 center-crop is extracted.

The training was conducted on a high-performance cluster at RWTH Aachen University. We adopted a ResNet50 model pre-trained on ImageNet. The binary cross-entropy loss was used for our binary classification tasks. Training spans 50 epochs. The model employs the SGD optimizer with a learning rate of 0.01 and momentum of 0.9. A learning rate scheduler reduces the learning rate by a factor of 10 every 7 epochs, aiding in precise model tuning as training progresses (reduce by < 1 = increase). Model performance was evaluated based on accuracy and ROC-AUC metrics, with periodic checks to save the best-performing model based on the highest ROC-AUC achieved. Predictions were made on the test set using these best models, and the predictions were evaluated and saved. The corresponding code can be found on GitHub (https://github.com/OMFSdigital/PAN-AI-X).

### Performance of Dentists

In addition, the test images were evaluated by 5 dentists/specialists (A.P., J.B., I.M., K.X., B.P.) with different levels of experience (dentist in first year to specialist in oral and maxillofacial surgery) to evaluate human performance. For this purpose, the 4,298 test images (2% margin) were randomly distributed among the investigators. Each dental image was then given a score between 0 (preserved) and 10 (extracted) to determine the the likelihood with which a human investigator would recommend a removal of the to.. The 2% margin was chosen to compare human performance to the DL model with the best performance. To avoid a learning effect between the annotation in the PANs and the scoring of the individual tooth images by the investigators, there was a 6-month time delay between initial annotation and scoring.

### Model Explainability

To explain the basis of the prediction of the AI models, CAMERAS [32] was used. It uses class activation mapping to help visualize the regions of the input image that are important for the model’s decision-making process (Figure 4, 5). In our case of binary classification where outcomes are extraction or preservation, CAMERAS highlights features based on the binary outcome. If the model predicts extraction, it highlights features leading to this decision; conversely, a prediction of preservation highlights or lacks features, indicating why the preservation is predicted. The intensity and frequency of these highlights can aid in interpreting model outputs, where more frequent or intense highlights correlates with a prediction with a higher probability.

### Statistical analysis

The statistical analysis was performed in Python (version 3.11.0) using the scikit-learn package (version 1.4.0). The performance of the AI classifiers and dentists was assessed by using the area under the curve of the receiver operating characteristic curve (ROC-AUC) and the precision-recall curve (PR-AUC). We then calculated the maximum Youden’s index for each ROC curve and acquired the optimal threshold for the corresponding model.

Metrics of accuracy, specificity, precision (syn. positive predictive value), and sensitivity (syn. recall) were calculated with the thresholds above The F1 score was calculated from precision and sensitivity. We used a set of thresholds of 0.3 and 0.7 to plot the confusion matrices with clinically relevant decisions, namely extraction, monitoring, and preservation.

## Results

### Patients

1,184 patients who met the criteria were selected in this study. The average age of patients was 50.0 years (range 11 – 99 years), with a standard deviation of 20.3 years. The gender ratio of the cohort was 61:39, with 722 males and 462 females. A total of 26,956 teeth were annotated in 1,184 PANs with bounding boxes and classified into preservation (21,797) and extraction (5,159). The prevalence of tooth extraction in our dataset was 19.1%, compared to the majority of 80.9% of preserved teeth. The demographic and clinical characteristics of patients are described in Table 1.

**Table 1.**
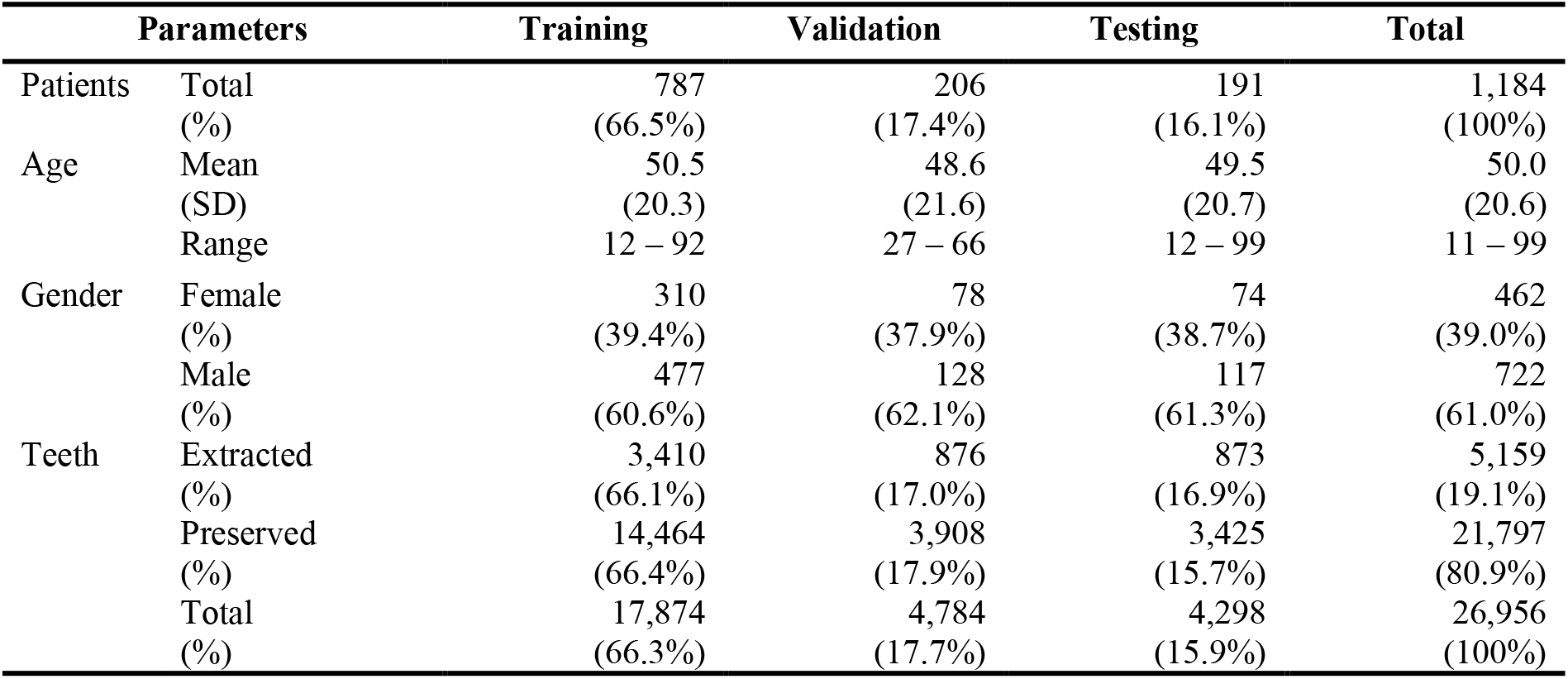
Demographic and dental characteristics of the patients and distribution across training, validation, and testing datasets.

### Performance of AI models

Eight different ResNet-50 models were trained on single tooth images with margin settings from -0.5% to 10%. The performance of models is summarized in Table 2 and Figure 2 based on the thresholds at the maximum Youden’s index. The model with 2% margin setting yielded the best results in both ROC-AUC (0.901) and PR-AUC (0.749). It also exhibited the best performance in all other metrics except for sensitivity. Shrinking of the bounding boxes (margin -0.5%) produced worse results in ROC-AUC and PR-AUC than the baseline (margin 0%). A general increase can be observed in both ROC-AUC and PR-AUC as the margin increases from -0.5% to 2%. Models with a 5% margin setting have achieved the highest sensitivity (0.835). However, increasing the margin further to 10% reduced both ROC-AUC and PR-AUC. In confusion matrices, with thresholds of 0.3 and 0.7 for monitoring, the 2% margin model had the least cases of false positive (53). The model with 3% margin had the highest accuracy (3455/4298).

**Table 2.** Performance at Youden’s index of AI models with different margin settings as well as human performance..

| Predictor | Margin | Accuracy | Sensitivity | Specificity | Precision | F1_Score | ROC-AUC | PR-AUC |
| --- | --- | --- | --- | --- | --- | --- | --- | --- |
| AI<br>(ResNet 50) | -0.5% | 0.775 | 0.700 | 0.794 | 0.464 | 0.558 | 0.815 | 0.590 |
|  | 0% | 0.768 | 0.781 | 0.765 | 0.459 | 0.578 | 0.852 | 0.657 |
|  | 0.5% | 0.778 | 0.772 | 0.780 | 0.472 | 0.586 | 0.852 | 0.650 |
|  | 1% | 0.813 | 0.716 | 0.837 | 0.529 | 0.608 | 0.864 | 0.693 |
|  | **2%** | **0.834** | 0.797 | **0.843** | **0.564** | **0.661** | **0.901** | **0.749** |
|  | 3% | 0.812 | 0.805 | 0.814 | 0.525 | 0.635 | 0.895 | 0.743 |
|  | 5% | 0.805 | **0.835** | 0.797 | 0.512 | 0.634 | 0.899 | 0.741 |
|  | 10% | 0.817 | 0.816 | 0.818 | 0.533 | 0.645 | 0.890 | 0.727 |
| Human<br>(Avg.) | 2% | 0.775 | 0.674 | 0.800 | 0.462 | 0.548 | 0.797 | 0.589 |

**Figure 2.**
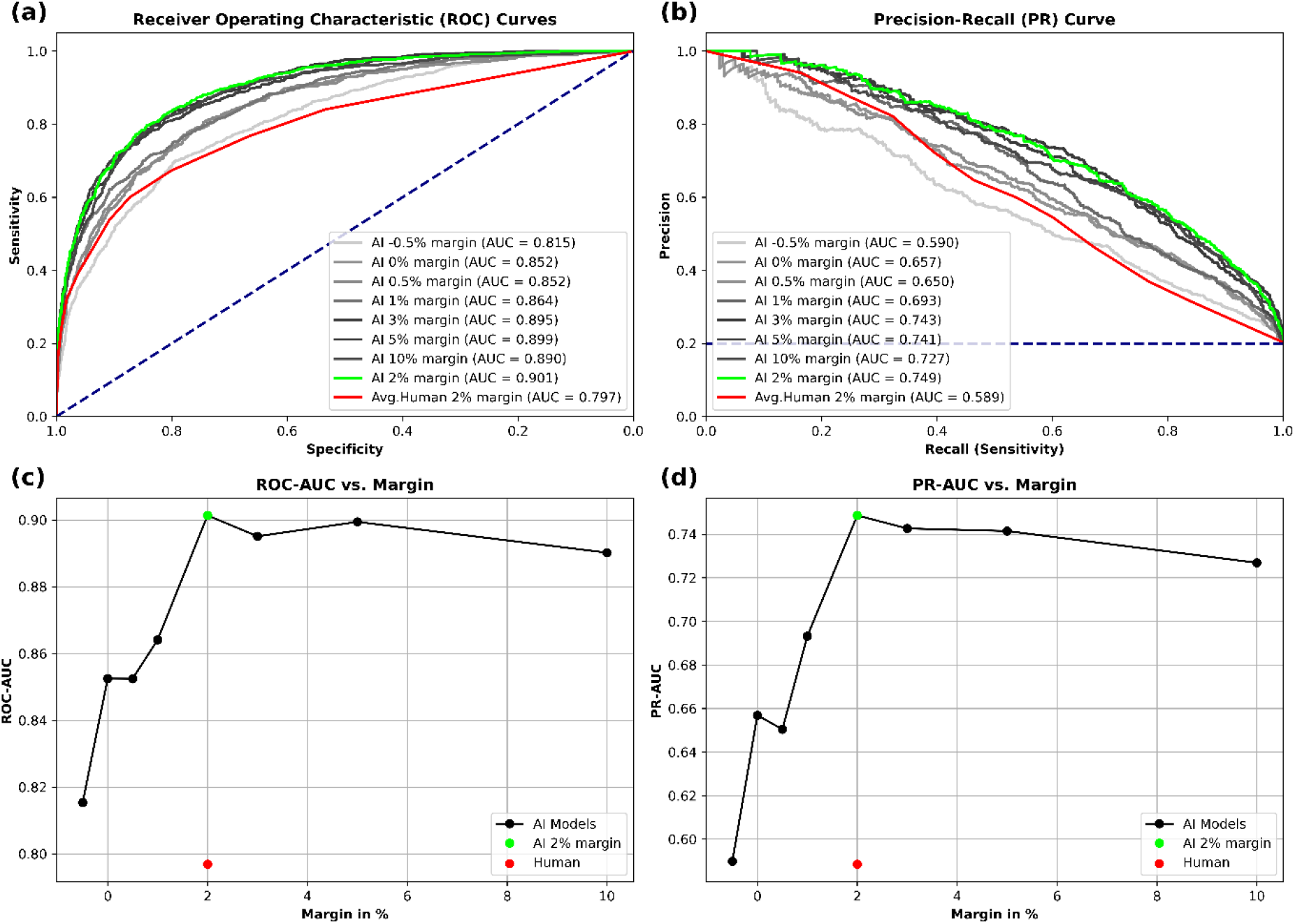
(**a**) ROC curves and (**b**) PPV-Sensitivity curves of models with different margin settings. The 2% margin model performed best in both ROC-AUC (0.901) and PR-AUC (0.749), the average human performance was ROC-AUC (0.797) and PR-AUC (0.589). Relationship between ROC-AUC and margins is displayed in (**c**). Relationship between PR-AP and margins is displayed in (**d**). A steep increase observed for both metrics from - 0.5% to 2% margin and slightly drop from 5% to 10% margin.

### Performance of Dentists

In contrast, the human assessment (average of 5 dentists/specialists) had a lower performance based on the 2% dental images compared to the AI models. The ROC-AUC was only 0.797 or PR-AUC of 0.589. This is also reflected by the confusion matrices where human have the most false positives (131) and lowest accuracy (3085/4298).

### Explainability

Figure 4 and 5 shows the activation map of the *extracted and preserved* predictions generated by CAMERAS with a 2% margin setting. In extraction cases, the model focused on the areas where roots are exposed in low density regions and crowns are buried in bone. In preservation cases, on the other hand, alveolar ridge and periapical regions were the most relevant.

## Discussion

In this study, to our knowledge, we present the first clinical prediction model using DL to make a recommendation about teeth extractions. The main results of the study are, 1) the best model achieved a ROC-AUC of 0.901 with a PR-AUC of 0.749; 2) outperforming dentists/specialists, who on average achieved a ROC-AUC of 0.797 with a PR-AUC of 0.589; 3) additional contextual information through wide margins around the tooth led to a better prediction; 5) the visual explainability of the prediction for tooth extraction or preservation was comprehensible.

Decision aids are a useful tool, for example in healthcare, to reduce the dentists’ workload, as suggestions calculated by algorithms can contribute to the final decision-making or diagnosis and significantly speed up this process [33]. Similarly, decision aids can be used as an objective perspective, especially in borderline cases where otherwise subjective approaches are applied by the clinicians alone [33, 34]. In this regard, work in the medical field has already been done on identifying pathologies in medical imaging like X-ray scans. One of the first applications used for detection was in 1995 to detect nodules in X-rays of the lungs [35]. Another object detection algorithm was developed to detect and classify several entities in chest X-rays like cardiomegaly, calcified granulomas, catheters, surgical instruments or thoracic vertebrae [36]. The emergence of convolutional neural networks / DL more than a decade ago opened up completely new possibilities [37].

One recent application is described by Yoo et al. who proposed a DL model (VGG16 pre-trained on ImageNet) to predict the difficulty of extracting a mandibular third molar from PANs [38]. The model was trained to predict the difficulty of mandibular third molar extraction in terms of depth, ramal relationship, and angulation. The accuracies of the model for different difficulty parameters (depth, ramal relationship, angulation) were found to be 78.9%, 82.0%, and 90.2%, respectively. Yet the model was made to predict the difficulty rather than the necessity of the extraction.

In our study, we used a residual neural network (ResNet-50) pretrained on ImageNet for the development of our clinical prediction model. Compared to other convolutional neural networks, a ResNet is characterized by so-called residual skip connections, which add inputs to outputs of small blocks of layers in the network. These skip connections improve the gradient flow during training and significantly improve the performance of very deep networks [39]. An outstanding strength of our model was its ability to classify teeth not worthy of preservation across multiple indications, such as extractions for orthodontic space, misplaced wisdom teeth, caries-destroyed teeth, periodontally compromised teeth or teeth from mixed dentition. Equally noteworthy was the reliable classification even in radiographs with more difficult classification conditions, such as anatomical superimposition effects.

Yet, evidence-based medicine encourages decisions based on patient-specific clinical evidence. However, DL models often provide blunt predictions without any explanation [40]. This results in a low acceptance among practitioners of these predictions due to the lack of visible evidence [29]. To address this problem, class activation map offers a solution to visualize and highlight the critical area of the image where the predictions are made [41, 32]. In the case of the caries classification task in the study of Vinayahalingam et al., areas that leads to the classification by DL model were be highlighted [42]. Such visual prompts can then correlate with established dental knowledge of the practitioners, which in turn explains the classification or recommendations.

We used CAMERAS, which, in contrast to methods such as GCAM or NormGrad, provides high-resolution mapping for ResNet and, thus, new insights into the explainability of DL methods [32]. The explainability can be illustrated using the examples of extracted teeth (Figure 4) and preserved teeth (Figure 5), including their prediction probability. In the case of healthy teeth, for example, this leads to activation of the bone, whereas in the case of root remnants this leads directly to the root itself. In addition to the recommendation, this activation map could also be offered directly to the dentist.

Interestingly, however, it can also be seen that due to the additional context information provided by the extended margin (2%) in Figures 4 and 5, neighboring root residues are also included in the classification and may possibly lead to a misclassification. This could be remedied in the future by more modern architectures that consider the entire PAN instead of individual image sections with a tooth and the adjacent bone.

Besides these technical aspects, the question arises as to how such a model could be translated into practice. An important challenge is that DL models fall under regulatory requirements such as FDA / Medical Device Regulation (MDR) as medical software. This means that the models developed in research cannot simply be applied in clinical encounters [43]. An important step here would be the external validation of the developed model [44]. At our department, the prevalence of tooth extraction was 19.1% (Table 1). This is influenced by the present population with its socioeconomic status, but certainly to some extent also to the treating specialty (conservative dentistry, prosthodontics, orthodontics, oral and maxillofacial surgery) has an impact that cannot be dismissed out of hand, as well as the pre-selection of cases. This could represent a bias if the model is applied elsewhere. On the other hand, it could be argued that the reasons for tooth extraction are universal worldwide [3, 45]. Periapical radiolucency or deep caries are not treated much differently around the world.

Clinical prediction models such as ours usually divide cases into two treatment recommendations based on a single threshold (perceive / extract). For an actual application scenario, however, the question of design is particularly crucial for optimal clinical usefulness [46]. This could involve dividing teeth into three groups based on two thresholds. Using a low threshold (with a high negative predictive value) to distinguish teeth that are definitely worth preserving from suspect teeth. Another higher threshold (with a high positive predictive value) could separate suspect teeth from definitely not preservable ones. The suspect teeth could then be monitored closely, while the healthy teeth would be ignored, and the decayed teeth would be extracted. An example for this approach is shown in Figure 3.

**Figure 3.**
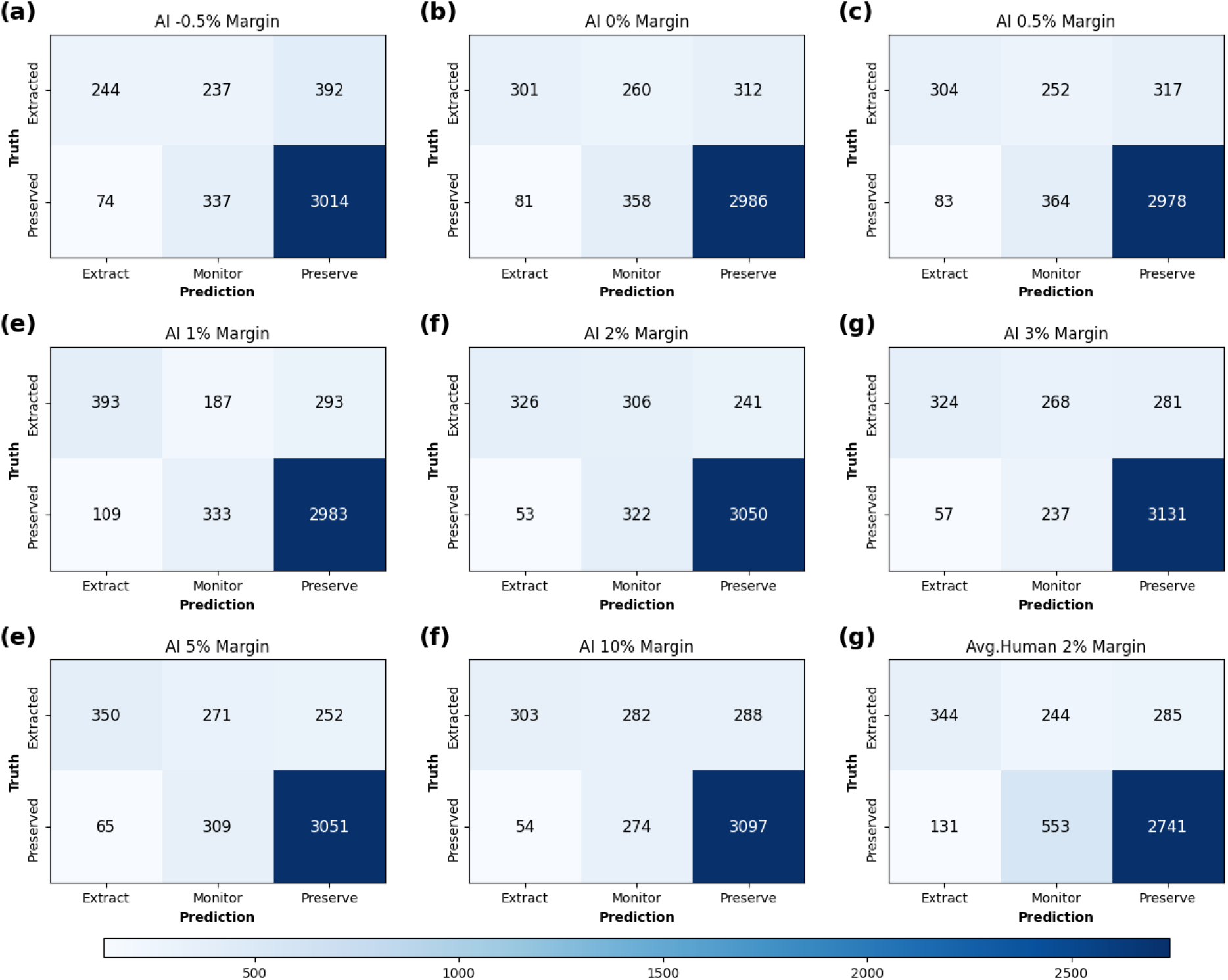
Confusion matrices showing prediction results. The results from AI models **(a)∼(f)** and dentists **(g)** with different margins were split into 3 decisions, namely extraction, monitoring, and preservation. Teeth with prediction probabilities from 0.3 to 0.7 were recommended to “Monitor”. Teeth with prediction probabilities below 0.3 were recommended to “Extract” while above 0.7 to “Preserve”. True labels were marked in y-axis.

**Figure 4.**
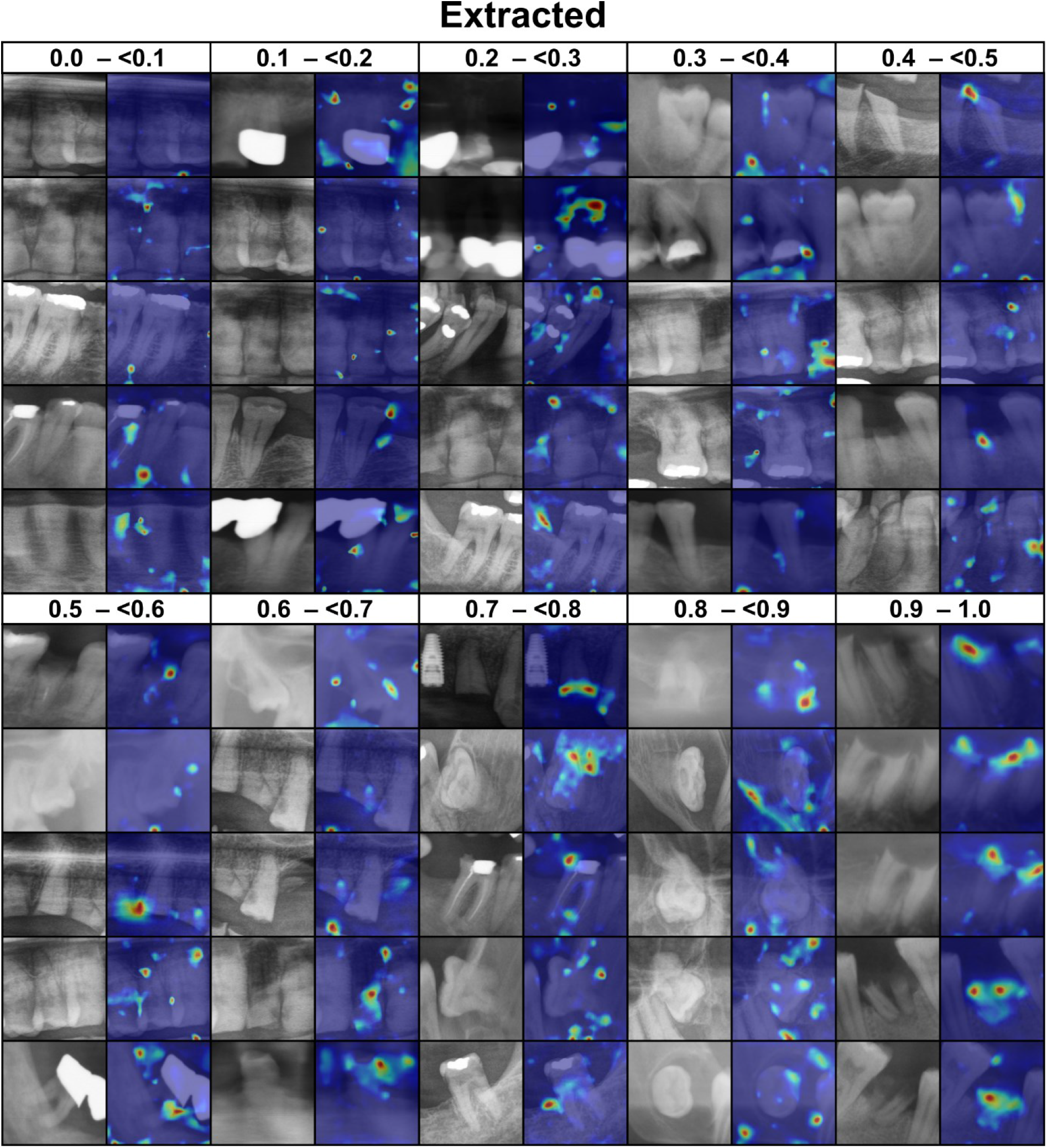
Activation gradient heatmap generated by CAMERAS for extracted teeth with a margin of 2%. The probability (0 to 1, where 0 indicates preservation and 1 indicates extraction) of the prediction is shown in the first row. The left image in each column is the tooth image used for the prediction, the right image is the class activation mapping with CAMERAS. Blue indicates no activation and red indicates strong activation. Green and yellow are in between.

**Figure 5.**
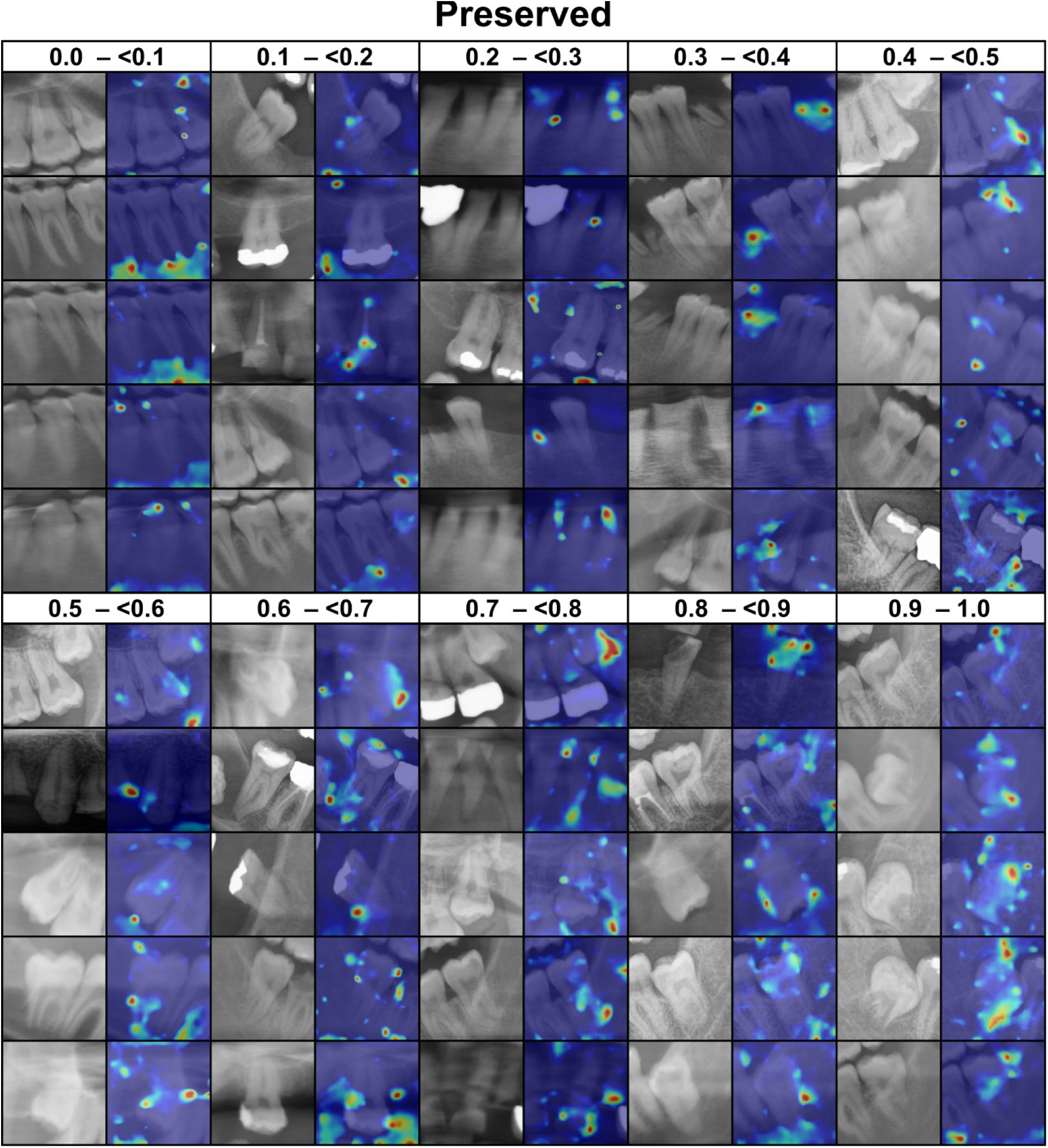
Activation gradient heatmap generated by CAMERAS for preserved teeth with a margin of 2%. The probability (0 to 1, where 0 indicates preservation and 1 indicates extraction) of the prediction is shown in the first row. The left image in each column is the tooth image used for the prediction, the right image is the class activation mapping with CAMERAS. Blue indicates no activation and red indicates strong activation. Green and yellow are in

However, a major limitation of our results is that our model does not include clinical information (pain, tooth vitality, course of disease, diagnosis). On the one hand, this is impressive because a high level of accuracy has been achieved despite the lack of any clinical information surpassing humans. Nevertheless, in a real clinical setting this information would be available and should be used. In the future, multimodal AI models could be used to process additional clinical information and improve prediction.

Another limitation is that there was a maximum period of 6 months between pre- and postoperative PAN. Usually, significant changes are visible during this period, but the causes for the extraction may not have been visible on the preoperative image used in some cases, but only shortly before the extraction itself (such as the involvement of teeth in a mandibular fracture).

## Conclusion

In summary, our study presented the first AI model to our knowledge to assist dentists/specialists in making tooth extraction decisions based on radiographs alone. The developed AI models outperform humans, with AI performance improving as contextual information increases. Future models may integrate clinical data. This study provides a good foundation for further research in this area. In the future, AI could help monitor at-risk teeth and reduce errors in indications for extraction. By providing a class activation map, clinicians could be able to understand and verify the AI decision.

## Data Availability

Code Availability Statement: All code was implemented in Python. The source code, including the model weights, is available on GitHub (https://github.com/OMFSdigital/PAN-AI-X).
Data Availability Statement: The data presented in this study are available upon reasonable request from the corresponding author.

https://github.com/OMFSdigital/PAN-AI-X

## Declaration

### Author Contributions

Conceptualization, B.P., I.M., and K.X.; methodology, I.M., L.S., J.R., A.H., B.P., and J.E.; software, L.S., I.M. and J.R.; validation, K.X., B.P., I.M., J.B., K.G. and A.P.; formal analysis, K.X., B.P., A.F., A.H., F.H. and D.T.; investigation, I.M., L.S., K.X. and B.P.; resources, B.P., F.H. and D.T.; data curation, I.M., K.X. and L.S.; writing—original draft preparation, K.X., B.P. and I.M.; writing—review and editing, B.P., K.X., I.M., L.S., J.B., K.G., A.P., J.R., A.F., A.H., J.E., F.H. and D.T.; visualization, B.P., L.S. and K.X.; supervision, B.P.; project administration, B.P.; All authors have read and agreed to the published version of the manuscript.

### Institutional Review Board Statement

The study approved by the Institutional Review Board (or Ethics Committee) of University Hospital RWTH Aachen (approval number EK 068/21, chairs: Prof. Dr. G. Schmalzing and PD Dr. R. Hausmann, approval date 25.02.2021).

### Informed Consent Statement

Informed consent was obtained from all subjects involved in the study.

### Code Availability Statement

All code was implemented in Python. The source code, including the model weights, is available on GitHub (https://github.com/OMFSdigital/PAN-AI-X).

### Data Availability Statement

The data presented in this study are available upon reasonable request from the corresponding author.

## Acknowledgments

Computations were performed with computing resources granted by RWTH Aachen University under project rwth1410.

